# Estimating the burden of COVID-19 pandemic on mortality, life expectancy and lifespan inequality in England and Wales: A population-level analysis

**DOI:** 10.1101/2020.07.16.20155077

**Authors:** José Manuel Aburto, Ridhi Kashyap, Jonas Schöley, Colin Angus, John Ermisch, Melinda C. Mills, Jennifer Beam Dowd

## Abstract

**Background:** Deaths directly linked to COVID-19 infection may be misclassified, and the pandemic may have indirectly affected other causes of death. To overcome these measurement challenges, we estimate the impact of the COVID-19 pandemic on mortality, life expectancy and lifespan inequality from week 10, when the first COVID-19 death was registered, to week 47 ending November 20, 2020 in England and Wales through an analysis of excess mortality.

**Methods:** We estimated age and sex-specific excess mortality risk and deaths above a baseline adjusted for seasonality with a systematic comparison of four different models using data from the Office for National Statistics. We additionally provide estimates of life expectancy at birth and lifespan inequality defined as the standard deviation in age at death.

**Results:** There have been 57,419 (95% Prediction Interval: 54,197, 60,752) excess deaths in the first 47 weeks of 2020, 55% of which occurred in men. Excess deaths increased sharply with age and men experienced elevated risks of death in all age groups. Life expectancy at birth dropped 0.9 and 1.2 years for females and males relative to the 2019 levels, respectively. Lifespan inequality also fell over the same period by five months for both sexes.

**Conclusion:** Quantifying excess deaths and their impact on life expectancy at birth provides a more comprehensive picture of the burden of COVID-19 on mortality. Whether mortality will return to -or even fall below-the baseline level remains to be seen as the pandemic continues to unfold and diverse interventions are put in place.

**Summary boxes:** *What is already known on this topic:* COVID-19 related deaths may be misclassified thereby inaccurately estimating the full impact of the pandemic on mortality. The pandemic may also have indirect effects on other causes due to changed behaviours, as well as the social and economic consequences resulting from its management. Excess mortality, the difference between observed deaths and what would have been expected in the absence of the pandemic, is a useful metric to quantify the overall impact of the pandemic on mortality and population health. Life expectancy at birth and lifespan inequality assess the cumulative impact of the pandemic on population health.

*What this study adds:* We examine death registration data from the Office for National Statistics from 2010 to week 47 (ending on November 20) in 2020 to quantify the impact of the COVID-19 pandemic on mortality in England and Wales thus far. We estimate excess mortality risk by age and sex, and quantify the impact of excess mortality risk on excess deaths, life expectancy and lifespan inequality. During weeks 10 through 47 of 2020, elevated mortality rates resulted in 57,419 additional deaths compared with baseline mortality. Life expectancy at birth for females and males over the 47 weeks of 2020 was 82.6 and 78.7 years, with 0.9 and 1.2 years of life lost relative to the year 2019. Lifespan inequality, a measure of the spread or variation in ages at death, declined due to the increase of mortality at older ages.

## Introduction

Estimating the number of deaths caused by the coronavirus (COVID-19) pandemic is an important challenge [1]. Insufficient testing capacity for SARS-CoV-2, the causative pathogen of coronavirus disease, especially during the early phases of the pandemic, misclassification of causes of death and definitional inconsistencies in counting COVID-19 deaths across different sources make the true toll of COVID-19 infections hard to estimate [2,3]. Moreover, interventions imposed during the pandemic may have indirectly affected other causes of death [4]. For example, both fear of COVID-19 and the overstretching of the healthcare system may have deterred care-seeking for both chronic and acute conditions, potentially increasing mortality from other, non-COVID, causes[5]. Similarly, restrictions might have decreased deaths from external causes such as road traffic accidents, or increased deaths from causes such as suicide.

To overcome these measurement challenges an alternative approach to estimate the mortality burden of COVID-19 is to quantify the number of deaths during the pandemic compared to a baseline level of what would have been expected if the pandemic had not occurred. This approach for estimating excess all-cause mortality has been widely used to quantify the mortality toll of previous epidemics such as influenza[6] or HIV[7], and has also begun to be applied for COVID-19 [1,8]. Excess mortality may be quantified in different ways and the “excess numbers of deaths’’ approach has been commonly used so far in England and Wales [4,8,9]. While this metric provides an important measure of the burden of the pandemic on a society, simply counting total excess deaths does not provide an understanding of the substantial variation by age and sex over time in elevated mortality risks[10,11], nor does it allow for a comparison of current mortality conditions with past conditions due to changes in population age structure over the period. Furthermore, excess deaths do not provide an understanding of the cumulative impact of the pandemic on summary indicators of population health such as life expectancy.

Life expectancy at birth is a commonly used age-standardised summary indicator of population health that expresses the average number of years a newborn would be expected to live given the death rates in a particular period[12]. While no individual would actually be expected to experience these death rates throughout their life, life expectancy provides a snapshot of the mortality profile of a population in a given period. Additionally, life expectancy is a comparable indicator of population health that does not require the arbitrary choice of a standard population as done with reported standardised death rates. Furthermore, as life expectancy is sensitive to the ages at which deaths occur and because it is comparable across time, it can shed additional light on the cumulative burden of a crisis such as COVID-19 on population health and enable comparisons with previous population health conditions. Lifespan inequality is another complementary indicator of population health with implications for public health planning, which has increasingly been reported in population health research [13–15]. While life expectancy is a measure of average mortality, lifespan inequality focuses on a second dimension of mortality, the variation in length of life between individuals in a population. It is possible for two populations to have the same life expectancy (i.e. average) with different levels of lifespan inequality because of the variation in the distribution of the ages of death. Thus, lifespan inequality provides a complementary perspective that reflects how unevenly population health improvements are shared within a population, which has important implications for health and social care planning. Trends over the twentieth-century from high-income countries, including England and Wales, show that as life expectancy and the modal age at death have increased, lifespan inequality has tended to decrease[16]. Nevertheless, the age dynamics driving improvement in each indicator are different. Reducing mortality at any age increases life expectancy. However for lifespan inequality to decrease when life expectancy is increasing, more lives need to be saved at younger than older ages, usually below life expectancy[13]. This compresses the distribution of deaths, making ages at death more similar.

We estimate all-cause excess deaths from week 10 (March 2-8), the week in which the first death attributable to COVID-19 was registered in England and Wales, to latest available data from week 47 of 2020 (ending November 20). Our work builds on existing estimates and approaches in three ways. First, we provide estimates disaggregated by age and sex, to highlight variations in excess deaths during the pandemic in England and Wales. Second, we develop refined model-based counterfactual estimates of excess deaths that better account for exposures and seasonal mortality patterns. We also systematically assess the sensitivity of excess deaths to different model-based estimates. Third, we provide estimates of life expectancy and lifespan inequality during the first 47 weeks of 2020 and compare them with previous mortality trends. By considering all three measures together: excess deaths, life expectancy and lifespan inequality, this study presents a comprehensive assessment of the mortality impacts of the COVID-19 pandemic thus far.

## Methods

### Data

We extracted all-cause death counts stratified by week of death registration and sex from 2010 to the week for which latest data were available (week 47 of 2020) from the ONS for England and Wales. While weekly mortality data are available by 5-year age groups for 2020, this level of disaggregation is not available for previous years. Therefore, we used 6 age-groups (0-14, 15-44, 45-64, 65-74, 75-84 and 85-older years of age) for modelling weekly deaths to harmonise weekly death data across 2010 to 2020, and used the 5-year age intervals for calculating life expectancy and lifespan inequality estimates for 2020. We also obtained population estimates from ONS from 2010 to 2019[17], and population projections for 2020[18]. As these projections represent the population at the mid-year point, we used standard interpolation techniques[19] to estimate weekly mean population by sex and age groups over the study period to use them as offset in the modelling strategy. Yearly death counts by 5-year age groups were used to calculate annual indicators[20] such as life expectancy and lifespan inequality. All analyses use publicly-available aggregated data. The population coverage of vital registration in England and Wales is complete. Between March and May 2020, 81.1% of all deaths and 86.5% of deaths involving COVID-19 were registered within 1 week of occurrence[21]. Death registration in this period witnessed increased efficiency compared to trends noted in previous years due to changes implemented in the Coronavirus Act 2020 [22,23]. Based on trends from past years, 92% of deaths are registered within 1 month of occurrence. As the extent of bias caused by registration delays is not properly understood, we do not attempt to implement any correction factors to minimize risks of over-correction and inflating our findings.

### Excess mortality

We estimated the baseline number of deaths in the absence of COVID-19 by fitting four different models. First, we fitted Generalized Additive Models assuming Negative Binomial and Poisson distributions of deaths during the period of study[24]. These models include a log-linear mortality trend by sex and age, smoothed effects for age and seasonality, and an interaction between age and seasonality (see Supplement section 1). The smoothed effects are stratified by sex. Third, we fitted a Generalized Poisson Linear Model adjusted for year-to-year seasonality[25], also known as extended Serfling model[26] (see Supplement Section 1). These previous models included indicator variables for systematic lags in death registration observed in weeks coinciding with holidays (weeks 1, 52 and 22 (Bank Holiday)). Finally, for our fourth model, we created a baseline by averaging the death rates observed in each week of the years 2015-2019 (see Supplement Figures 1 and 2).

**Figure 1.**
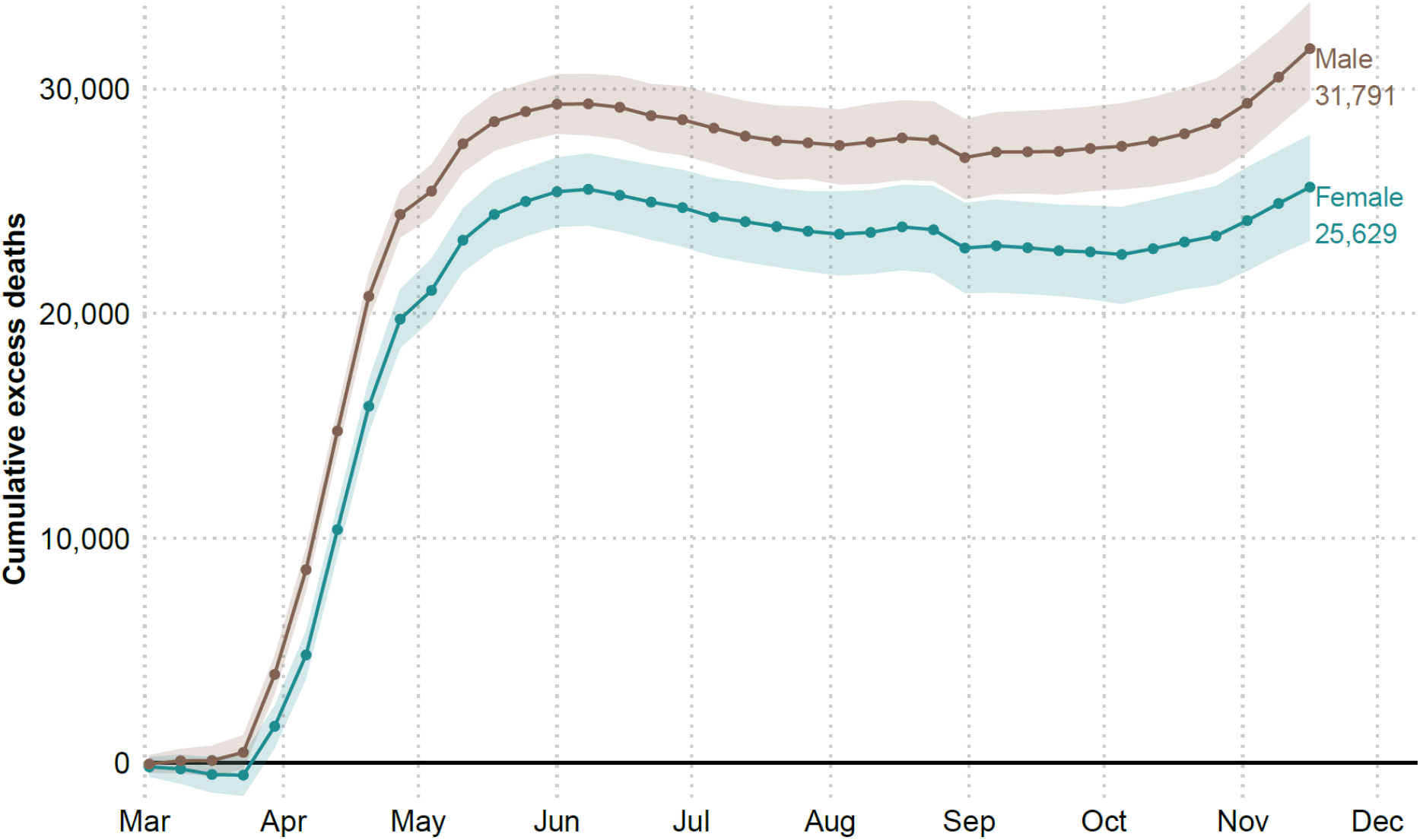
Cumulative excess deaths in England and Wales through the COVID-19 pandemic weeks 10-47 by sex. Shaded areas represent 95% prediction intervals. Excess deaths are defined as the total observed deaths subtracting the estimated baseline death count.

**Figure 2.**
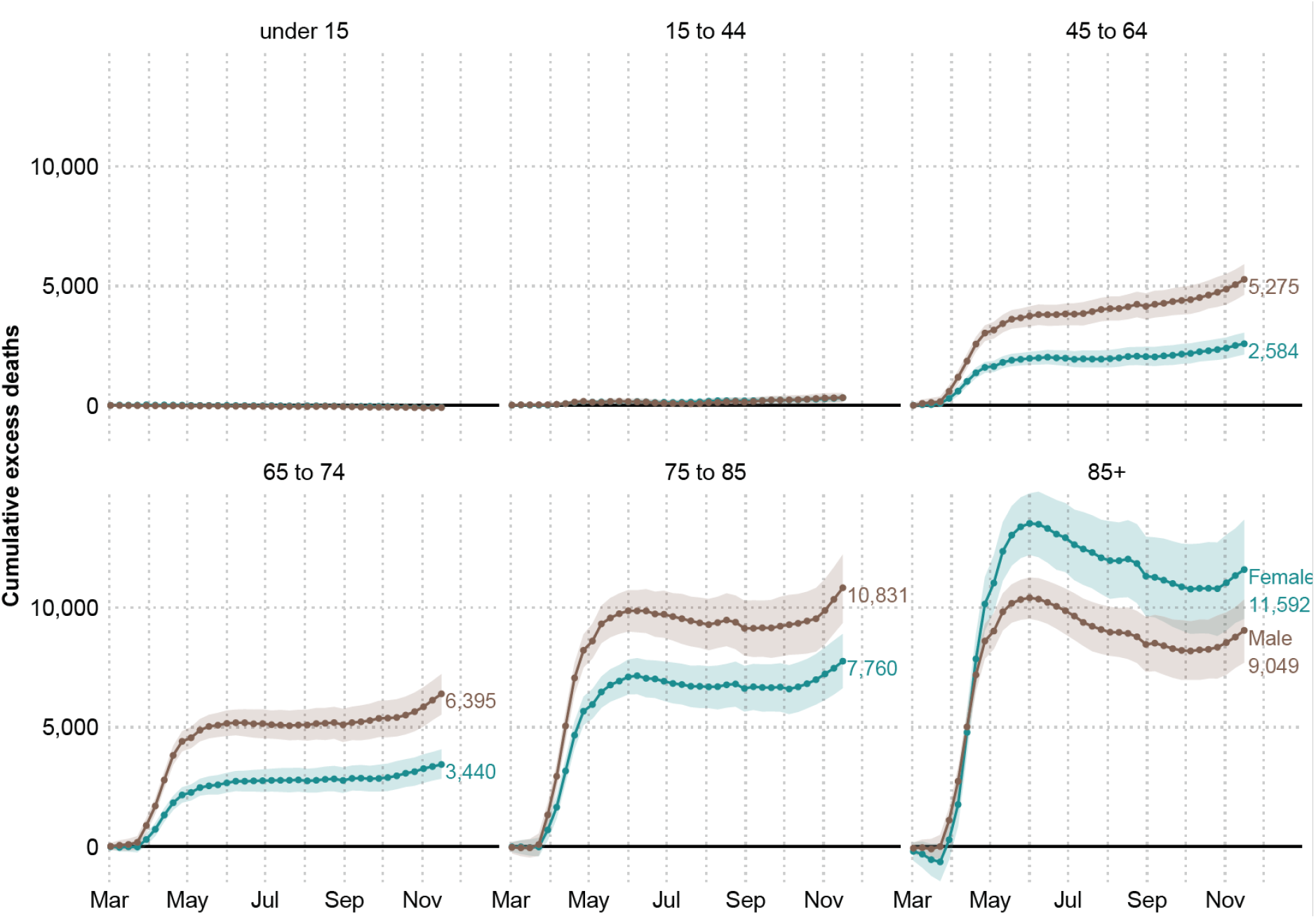
Cumulative excess deaths in England and Wales through the COVID-19 pandemic weeks 10-47 by sex and age groups. Shaded areas represent 95% prediction intervals. Excess deaths are defined as the total observed deaths subtracting the estimated baseline mortality.

**Figure 3.**
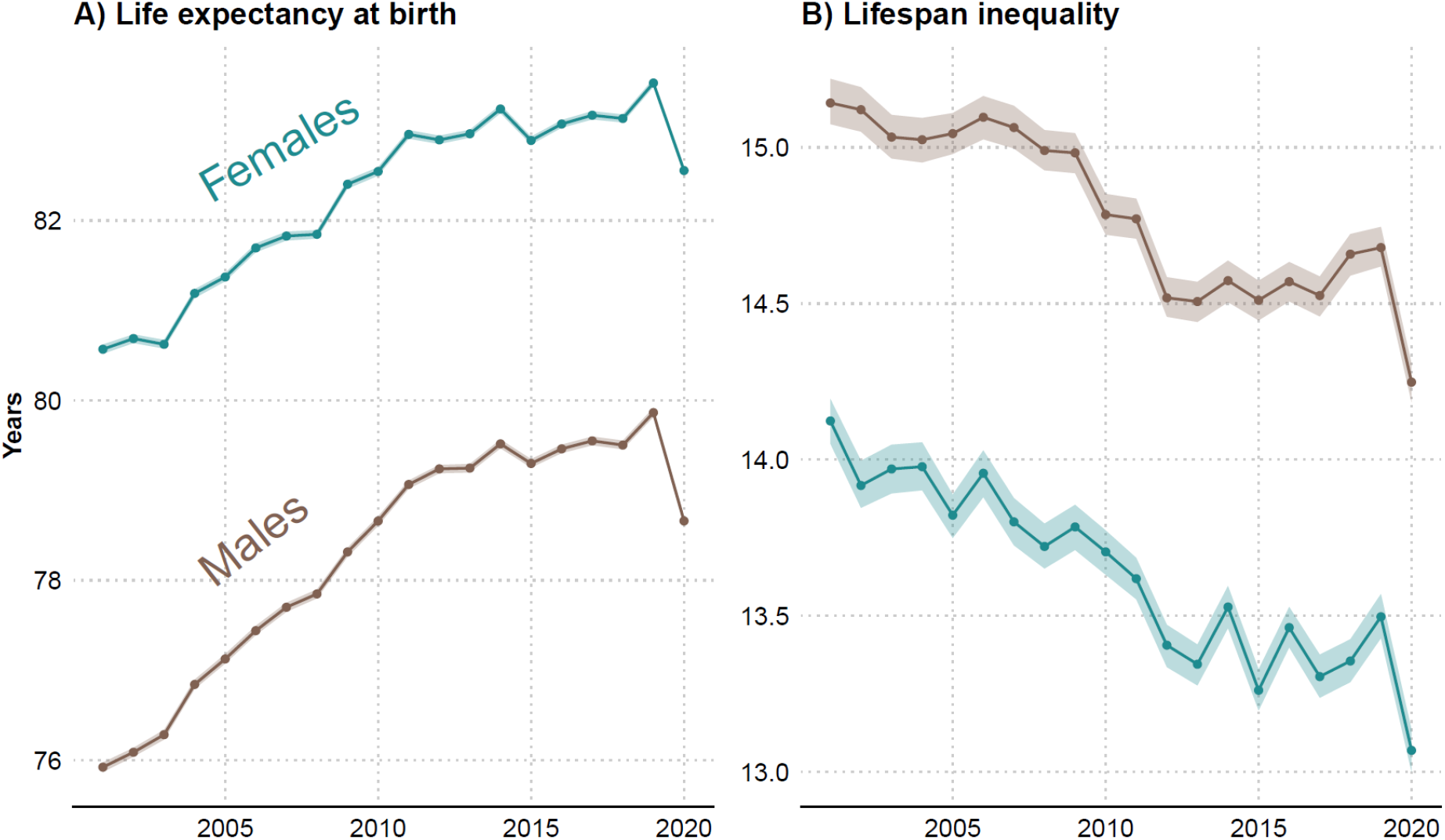
Life expectancy and lifespan inequality (standard deviation of ages at death) estimates for the periods 2001-2019, and for 2020 considering the first 47 weeks of the year by sex. Shaded areas represent 95% prediction intervals.

We fitted the models to the weekly deaths counts from January 4, 2010 to the week starting on March 2, 2020. This baseline was then projected forward until November 20, 2020 (week 47). Excess mortality is then defined as the observed weekly death count minus the baseline, summed across the pandemic period from March 2 (week 10) to November 20 (week 47), 2020. From this baseline, 95% predictive intervals were constructed by sampling death counts from Negative Binomial and Poisson distributions depending on the model’s underlying distribution.

We report excess death estimates from the negative binomial model in the main text but estimates comparing the different approaches are provided in the supplementary materials. This choice is based on out-of-sample predictive performance on past non-COVID weekly death counts.

### Demographic Methods

Life expectancy and lifespan inequality by sex were estimated using the yearly death counts and population estimates for the years preceding 2020 using standard demographic techniques [12], from which 95% predictive intervals were generated[27]. For the 47 weeks of 2020 for which data were available, death counts were aggregated over age groups and death rates were calculated using a proportionally adjusted mid-year population estimate.

### Code and Data availability

All analyses were carried out using R software[28]. All analysis scripts and data are available in a public repository and will be updated as more data become available[29].

## Results

### Estimates of excess deaths

The first death attributable to COVID-19 in England and Wales was registered in the week starting on March 2, 2020 (week 10). From that week until the end of week 47 on November 20, 2020, there were 436,102 registered deaths, from which an estimated 57,419 (54,197, 60,752) are excess mortality above the expected baseline (see Figure 1). This estimate represents a 15.1% (14.2, 16.2) increase in deaths compared to the expected level.

Death rates during the pandemic were consistently higher among males in all groups compared to females (see Supplement Figure 3). Male excess deaths accounted for 55.3% (31,791 deaths) of total excess deaths, compared to 44.6% (25,629 deaths) among females over the same period, despite the fact that females make up a larger fraction of the oldest old Between March 2 and November 20, male deaths exceeded the expectation by 16.8% (15.4, 18.0) and female death counts by 13.6% (12.2, 14.9). Cumulative excess deaths at the end of the first wave (week 26 ending in June 29) were 53,937 (95% Prediction Interval: 53,092, 54,746) followed by no excess mortality over the summer months, before an uptick that started in October 2020 when a second wave emerged and excess deaths began to rise again.

Disaggregating by age, we estimate no excess deaths among those younger than 15 years. The 15 to 44 year old age group accounted for 652 (395, 903) excess deaths (6.2% (3.6, 8.7) above the expected level). For older age groups excess deaths rose sharply (see Figure 2). The toll of the pandemic resulted in 7,859 (7,065, 8,645) and 9,835 (8814, 10,833) excess deaths among people between 45-64 and 64-74 years of age, respectively. These numbers are 17.6% (15.6, 19.7) and 16.0% (14.1, 17.9) above the baseline. The largest numbers of lives lost were estimated among the groups 75-85 and 85 and older, with 17.2% (15.3, 19.2) and 13.7% (11.9, 15.4) more deaths than expected. Among the former, 18,591 (16,845, 20,435), excess deaths were estimated, while among the oldest age group there were 20,641 (18,271, 22,916) deaths above the baseline. Note the larger number of female excess deaths in the 85+ group is due to there being 1.6 times more females in this age group compared to males. After a peak in excess deaths by June 2020, the 85+ age group saw somewhat lower than baseline mortality over the summer months, before an increasing trend in excess deaths emerged again in the second wave from October 2020. In contrast, for all other age groups, mortality remained at baseline over the summer months.

### Estimates of life expectancy and lifespan inequality

Female life expectancy at birth increased from 81.4 (81.3, 81.4) years in 2005 to 83.5 (83.5, 83.6) years in 2019 in England and Wales. Similarly, male life expectancy increased from 77.1 (77.1, 77.2) to 79.9 (79.8, 79.9) years in the same period. Using data from the first 47 weeks of 2020 yields an estimated life expectancy at birth of 82.6 (82.5, 82.6) and 78.7 (78.6, 78.7) for females and males, respectively, a reduction of 0.9 years for females and 1.2 years for males.

From 2005 to 2019, lifespan inequality declined slowly from 13.8 (13.7, 13.9) to 13.5 (13.4, 13.6) years for females and from 15.0 (15.0, 15.1) to 14.7 (14.6, 14.7) years for males. Over the first 47 weeks of 2020, we estimate that lifespan inequality fell sharply to 13.1 (13.0, 13.1) and 14.2 (14.2, 14.3) years for females and males, respectively, corresponding to a reduction of nearly five months for both sexes.

## Sensitivity analysis

We performed several sensitivity analyses. Firstly, we refitted the seasonal baseline without including the first 9 weeks of 2020. This adjustment did not have major effects on our estimates and by taking the first 9 weeks into account we aligned our predictions with the observed trend at the beginning of the year. Our four models produce central estimates of the number of excess deaths between 49,056 and 57,419 depending on the choice of the model and its assumptions, but do not substantively affect the pattern of our results. We note that excess deaths derived from the baselines estimated from both the Generalized Additive Models and Generalized Linear Models indicated a higher magnitude of excess deaths than those using average mortality rates from the preceding five-years as the baseline. For full details see Supplement Tables 1 and 2. In addition, we also estimated life expectancy using a piecewise constant hazard model and the results did not change.

## Discussion

Excess deaths during the first 47 weeks of the year 2020 shed light on the cumulative burden of the COVID-19 pandemic in England and Wales. While several European countries have experienced substantially increased mortality over the course of the pandemic, data at hand suggest that England and Wales are amongst the worst performers in terms of excess deaths, especially in the working-age group 15 to 64[30]. We estimated 57,419 (54,197, 60,752) premature deaths due to the pandemic. Our estimate is based on a systematic comparison of different approaches to estimating a mortality baseline from which excess is derived, and relies on a refined model that accounts for changes in population ageing and seasonality. The toll of the pandemic had unequal impacts by age and sex in Europe and other regions[10,31,32]. Similarly for England and Wales and consistent with other work[8], we found that excess mortality varied between sexes, with males accounting for 55% of excess deaths. Excess deaths increased sharply over age and male deaths were estimated to exceed females in all age groups, with the exception of those above age 85. This is explained by the population composition of England and Wales where more females survive to higher ages. Accounting for this compositional effect, death rates during 2020 were higher among men in all ages groups (see Supplement Figure 3).

According to the ONS, between March 1st and June 30th, 2020 there were 50,335 deaths involving COVID-19, 46,736 (93%) of which assigned COVID-19 as the underlying cause of death based on information noted on the death certificate[42]. A sizable fraction of our estimate for excess deaths over the first wave of the pandemic is thus likely to be directly linked to COVID-19. Based on preliminary cause of death analysis of other (non-COVID) causes by the ONS, deaths occurring from Alzheimer disease and dementia, ischemic heart disease, cerebrovascular diseases, influenza and pneumonia and ‘symptoms signs and ill-defined conditions’ category were all higher between March and May 2020 [22]. Together Alzheimer and ‘symptom signs and ill-defined conditions’ experienced the largest increases in magnitudes compared to previous years, and deaths occurring from asthma and diabetes at home also increased[22]. These preliminary cause-of-death patterns suggest that a significant fraction of the unexplained excess mortality over the first wave of the pandemic may also be attributable to undiagnosed COVID-19. As more detailed cause-of-death data become available over the coming months, future research should seek to develop methods to disentangle excess deaths attributable to COVID-19 versus those arising indirectly due to effects such as reduced care for other conditions.

For the latter half of the year, in the period from June 15 to the end of August (weeks 25 to 36), our estimates showed no excess mortality in most weeks for those under 85 before the emergence of a second wave of excess mortality from October. The lower than baseline mortality observed in the summer months of the 85+ age groups suggest potential mortality displacement effects, i.e., that some deaths were brought forward in this age group due to the pandemic, although the numbers far from compensated for first wave deaths even at these oldest ages. However, no similar signs of mortality displacement due to lower than baseline mortality were visible for the other age groups over the summer. As these estimates are based on deaths registered so far, it is too early to clarify the contributions of mortality displacement to excess mortality observed during the pandemic, and its impacts on post-crisis mortality levels.

Life expectancy in England and Wales had been steadily improving for 50 years before stagnating in the past decade [33,34]. We have provided estimates of life expectancy for 2019 and for 2020 based on mortality data until week 47 which show that life expectancy dropped a staggering 0.9 and 1.2 years for females and males respectively between these years. Moreover, our estimates for life expectancy fall 0.7 and 1.1 years below the official projected life expectancy in 2020 for females and males[35], respectively. To put this into perspective, male and female life expectancy regressed to the levels of 2010. It is likely that our estimates of excess deaths and life expectancy losses until this period are underestimated, as these estimates are based on deaths registered so far, a small fraction of which may have experienced registration delays[21]. Recent evidence suggests that reversals and stagnation in life expectancy amongst developed countries are usually a result of mid-life mortality crises [34]. In contrast, life expectancy losses during the pandemic have come about from sharp increases in older age mortality in both sexes.

Historically, life expectancy increases have been accompanied by reductions in lifespan inequality due to mortality improvements at younger ages, although more recently studies have found that life expectancy improvements can occur even without accompanying reductions in lifespan inequality [13]. Our results strikingly show a third, previously undocumented pattern of life expectancy and lifespan inequality change, with both decreasing concurrently due to the unique nature of the mortality stress triggered by the COVID-19 pandemic. In contrast with previous influenza pandemics such as the 1918-20 Spanish flu that primarily affected the young[36], or the 1957 pandemic that affected both the young and old[37], the mortality impact of the COVID-19 pandemic has primarily affected older age groups. Within a broader context of population health in which mortality is now largely concentrated at older ages, the elevated excess death rates at older age groups observed during the COVID-19 pandemic so far has reduced life expectancy. However, the disproportionate shift in the distribution of ages at death to older age groups made ages at death more similar thereby reducing variation but at the expense of increasing overall average mortality. As a result of these dynamics, life expectancy and lifespan inequality moved in the same direction.

Looking forward, it is unclear if life expectancy will return to baseline levels rapidly, and even if/when it recovers, how mortality will be different. The prospect of vaccination being likely in the near future suggests a potential for the rapid recovery of life expectancy, although this will depend on the rollout speed, coverage and efficacy of the vaccine[38]. In contrast, the combination of potential scarring effects of Covid-19, such as the long term consequences of the disease on individuals’ health[39], the implications of lockdown and non-pharmaceutical interventions on behaviours and mental health [40], cancer treatment delay associated with increased mortality[41], and the unequal impact of Covid-19 across subgroups by age, sex, ethnicity, SES and regions[42,43], could create an unseen mortality profile that maintains life expectancy at lower levels beyond the short-term into the medium-term.

## Data Availability

All data and scripts are available at a public repository.

https://doi.org/10.5281/zenodo.3946492

## Ethical approval

This article used aggregated, fully anonymized, publicly available data. Therefore no ethics approval is needed.

## Data sharing statement

This analysis used publicly available data. All data and scripts are available at https://doi.org/10.5281/zenodo.3946492

## Contributors

JMA, RK, JS, and JBD contributed to the design of the study. JMA, CA and RK drafted the manuscript. JS and JMA performed the statistical analysis. All authors contributed to interpretation of data, revised the article critically for important intellectual content, and approved the final version of the manuscript. JMA and RK, the corresponding authors, attest that all listed authors meet authorship criteria and that no others meeting the criteria have been omitted.

