## Supplement for "Estimating the burden of COVID-19 pandemic on mortality, life expectancy and lifespan inequality in England and Wales: A population-level analysis"

**Authors:** José Manuel Aburto, Newton fellow[1,2], Ridhi Kashyap, associate professor[1], Jonas Schöley, postdoctoral fellow[2], Colin Angus, senior research fellow[3], John Ermisch, professor[1], Melinda C. Mills, professor[1], Jennifer Beam Dowd, associate professor[1]

**Affiliations:**
[1] Leverhulme Centre for Demographic Science, Department of Sociology and Nuffield College, University of Oxford, 42-43 Park End Street, OX1 1JD Oxford, UK.

[2] Interdisciplinary Centre on Population Dynamics, University of Southern Denmark, Odense 5000, Denmark.

[3] School of Health and Related Research, University of Sheffield, Regent Court, Regent Street, S1 4DA Sheffield, UK

**Correspondence to:**
José Manuel Aburto
42-43 Park End Street, OX1 1JD Oxford, UK.

ORCID: 0000-0002-2926-6879

OR

Ridhi Kashyap
Nuffield College, New Road, Oxford OX1 1NF

ORCID: 0000-0003-0615-2868

**Section 1. Estimation of the baseline mortality risk using 4 different approaches using training data from 2010 to week 10 of 2020 by age and sex.**

1. Generalized Additive Model regression on weekly death counts assuming a Negative Binomial distribution to account for overdispersion of deaths during the period we study[1]. The model includes
    a. log-linear long-term mortality trends stratified by age group and sex,
    b. smoothed effects for mortality over age and seasonality stratified by sex,
    c. a smoothed interaction for mortality between age and seasonality stratified by sex, and
    d. age and sex specific effects for "special weeks", i.e. the first and last week of a year and week 21 where holidays cause registration delays,
    e. logged exposure times as offset.

    The structure of the model is as follows:

$$log\left(E(Y_i)\right) = \beta_0 + \beta_1 sex_i + \beta_2 agegroup_i + \beta_3 specialweek_i + \beta_4 time_i + \beta_5 sex_i * agegroup_i + \beta_6 time_i * sex_i + \beta_7 time_i * agegroup_i + \beta_8 time_i * sex_i * agegroup_i + \beta_9 specialweek_i * sex_i + \beta_{10} specialweek_i * agegroup_i + \beta_{11} specialweek_i * sex_i * agegroup_i + f_1(age_i | sex_i) + f_2(weekofyear_i | sex_i) + f_3(weekofyear_i, age_i | sex_i) + log(\theta_i)$$

    Where $E(Y_i)$ are the expected deaths in a given week and population stratum, $\theta_i$ are the population exposures, and $f$ are smoothed functions of continuous covariates. $f_1$ is a penalized spline for the age effect with a dimension 6 basis, $f_2$is a penalized cyclic spline over week of year for the seasonality effect with a dimension 8 basis and $f_3$is a smoothed interaction of age and seasonality.

2. The second approach is a Generalized Additive Model assuming a Poisson distribution with the same structure as above.

3. The third approach is a Generalized Linear Model regression on weekly deaths assuming a Poisson distribution and featuring trigonometric terms for the seasonal effect. These so-called Serfling models [2,3] are used to estimate baseline mortality during influenza epidemics. The basic structure of the model is as follows:

$$log\left(E(Y_i)\right) = \beta_0 + \beta_1 time_i + \gamma_2 sin\left(\frac{2\pi\ weekofyear_i}{52}\right) + \gamma_3 cos\left(\frac{2\pi\ weekofyear_i}{52}\right) + \gamma_4 sin\left(\frac{2\pi\ weekofyear_i}{26}\right) + \gamma_5 cos\left(\frac{2\pi\ weekofyear_i}{26}\right) + log(\theta_i),$$

    where all the terms are fully interacted with age and sex and "special weeks" are included in the same way as in the Generalized Additive models.

4. We constructed an empirical baseline mortality based on the average mortality rate over the previous five years 2015-19 within each week and stratum. The associated deaths from this approach result from multiplying the average death rates by the population exposed to the risk.

5. **Excess deaths produced with different models.**

Table 1. Total excess deaths by the end of week 47 estimated with 4 different models with 95% prediction intervals in England and Wales.

| | Female | | | Male | | | Total | | |
|---|---|---|---|---|---|---|---|---|---|
| **Model** | **Excess** | **.05 PI** | **.95 PI** | **Excess** | **.05 PI** | **.95 PI** | **Excess** | **.05 PI** | **.95 PI** |
| GAM Negative Binomial | 25,629 | 23,244 | 27,954 | 31,791 | 29,539 | 33,853 | 57,419 | 54,197 | 60,752 |
| GAM Poisson | 25,597 | 24,721 | 26,487 | 31,724 | 30,893 | 32,667 | 57,321 | 56,044 | 58,538 |
| GLM Poisson (Serfling) | 25,341 | 24,464 | 26,188 | 31,463 | 30,605 | 32,337 | 56,804 | 55,605 | 57,933 |
| Average mortality | 22,087 | 21,269 | 22,969 | 26,969 | 26,110 | 27,801 | 49,056 | 47,867 | 50,319 |

Table 2. Total excess deaths by the end of week 47 estimated with 4 different models by age and sex with 95% predictive intervals in England and Wales.

| | | **Female** | | | **Male** | | |
|---|---|---|---|---|---|---|---|
| **Model** | **Age group** | **Excess** | **.05 PI** | **.95 PI** | **Excess** | **.05 PI** | **.95 PI** |
| GAM Negative Binomial | 0 | -72 | -142 | -3 | -88 | -161 | -14 |
| | 15 | 324 | 172 | 467 | 329 | 109 | 527 |
| | 45 | 2,584 | 2,135 | 3,044 | 5,275 | 4,634 | 5,907 |
| | 65 | 3,440 | 2,845 | 4,070 | 6,395 | 5,531 | 7,221 |
| | 75 | 7,760 | 6,636 | 8,903 | 10,831 | 9,365 | 12,223 |
| | 85 | 11,592 | 9,536 | 13,674 | 9,049 | 7,704 | 10,338 |
| GAM Poisson | 0 | -76 | -139 | -12 | -90 | -164 | -15 |
| | 15 | 328 | 201 | 452 | 334 | 166 | 507 |
| | 45 | 2,586 | 2,337 | 2,830 | 5,269 | 4,971 | 5,565 |
| | 65 | 3,407 | 3,110 | 3,735 | 6,283 | 5,923 | 6,662 |
| | 75 | 7,719 | 7,269 | 8,181 | 10,770 | 10,296 | 11,235 |
| | 85 | 11,634 | 11,063 | 12,264 | 9,159 | 8,684 | 9,624 |
| GLM Poisson (Serfling) | 0 | -75 | -139 | -8 | -89 | -160 | -18 |
| | 15 | 322 | 196 | 441 | 331 | 162 | 491 |
| | 45 | 2,583 | 2,313 | 2,841 | 5,248 | 4,939 | 5,581 |
| | 65 | 3,332 | 3,029 | 3,661 | 6,225 | 5,861 | 6,597 |
| | 75 | 7,679 | 7,242 | 8,120 | 10,665 | 10,208 | 11,142 |
| | 85 | 11,500 | 10,916 | 12,081 | 9,083 | 8,606 | 9,535 |
| Average mortality | 0 | -187 | -253 | -122 | -234 | -308 | -156 |
| | 15 | 252 | 137 | 369 | 176 | 10 | 343 |
| | 45 | 2,146 | 1,863 | 2,418 | 4,644 | 4,324 | 4,976 |
| | 65 | 2,767 | 2,459 | 3,083 | 5,120 | 4,761 | 5,480 |
| | 75 | 5,676 | 5,233 | 6,103 | 8,495 | 8,008 | 8,962 |
| | 85 | 11,433 | 10,843 | 12,066 | 8,768 | 8,275 | 9,256 |

**Figure 1. Expected (lines) vs. observed deaths (points) counts based on the 4 approaches described above for males by age groups (rows) 0-14, 15-44, 45-64, 65-74, 75-84 and 85-older years of age. Shaded areas indicate 95% prediction intervals.**

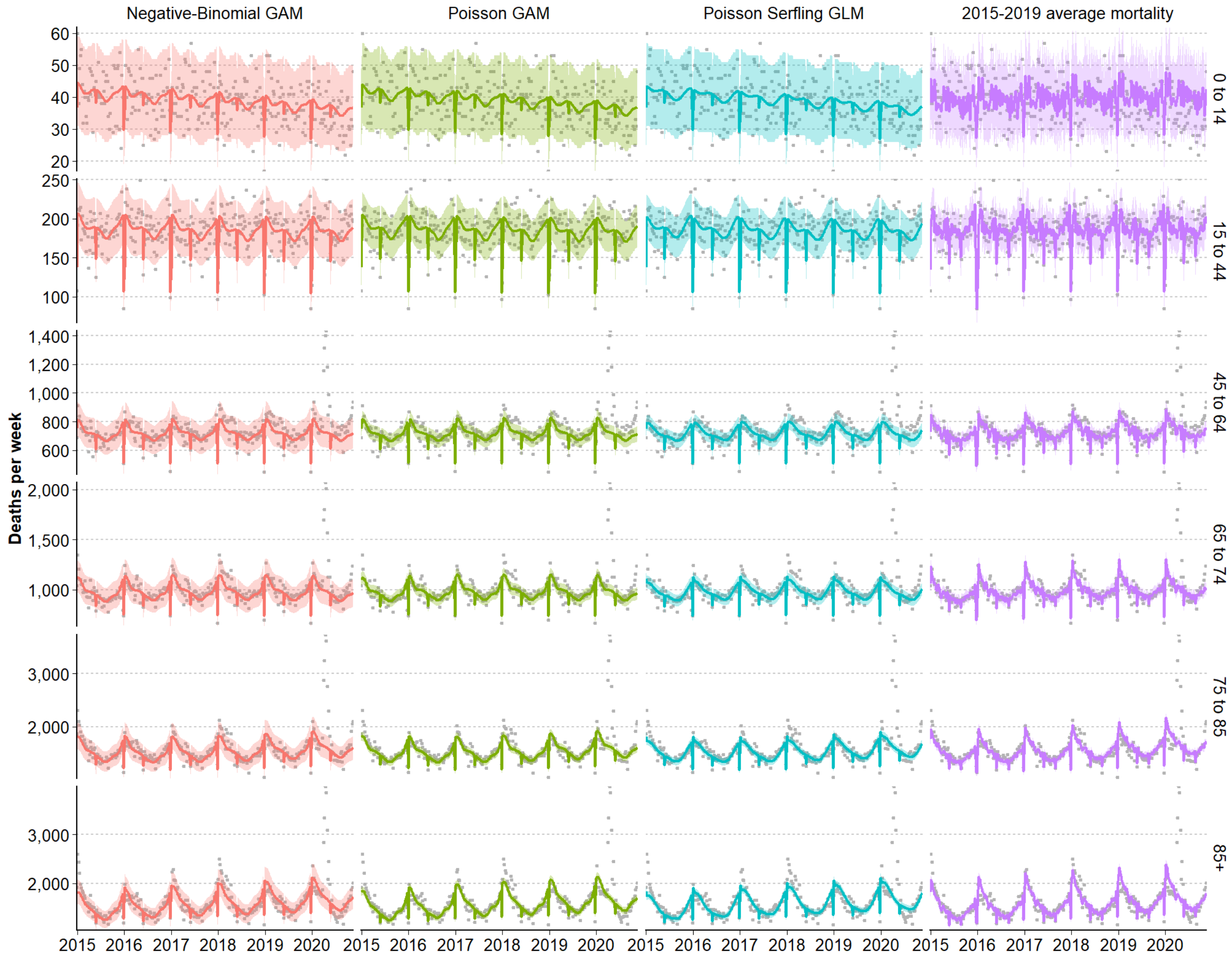

**Figure 2. Expected (lines) vs. observed deaths (points) counts based on the 4 approaches described above for females by age groups (rows) 0-14, 15-44, 45-64, 65-74, 75-84 and 85-older years of age. Shaded areas indicate 95% prediction intervals.**

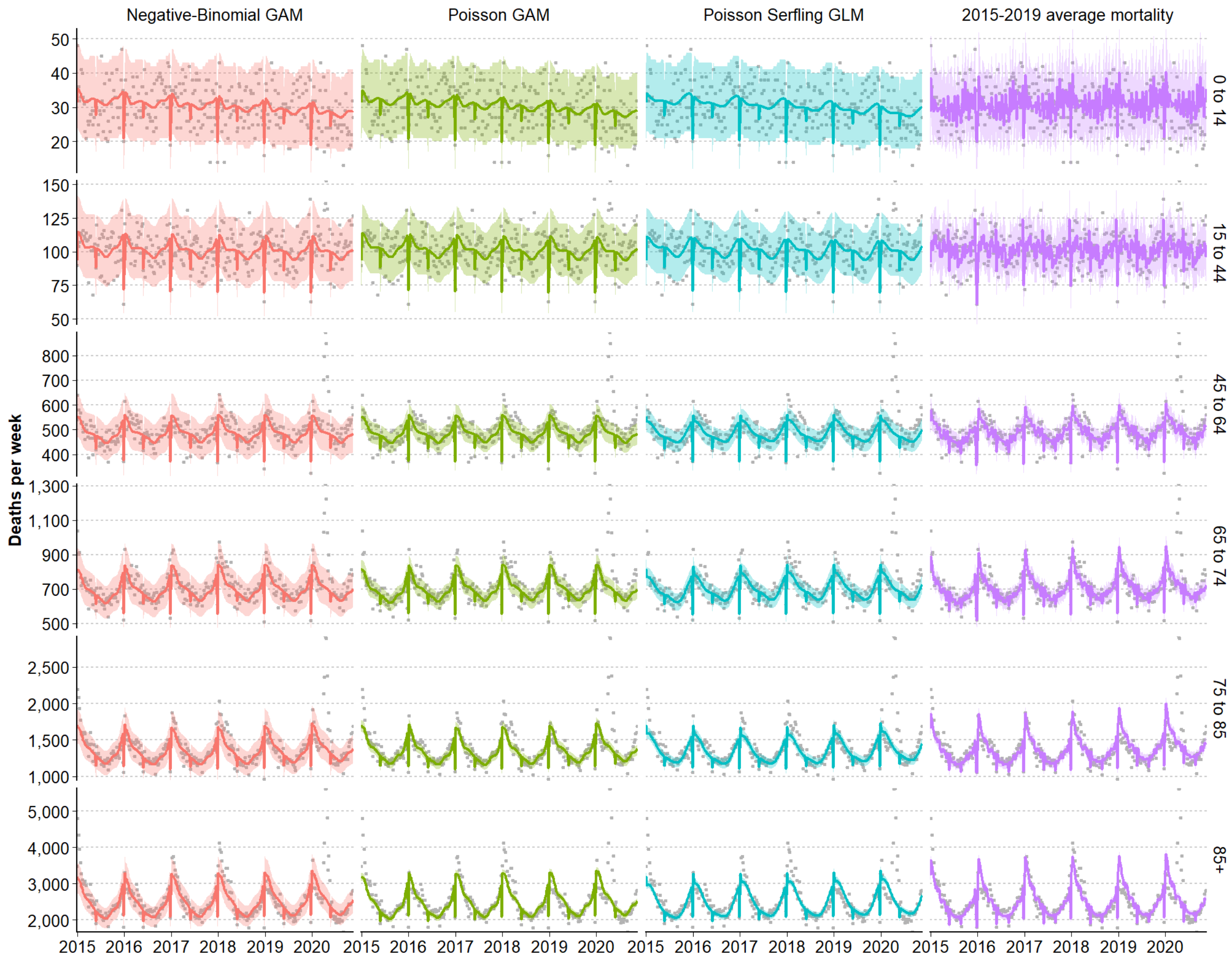

**Figure 3. Sex ratio males/females of death rates during the course of the pandemic by age groups (rows) 0-14, 15-44, 45-64, 65-74, 75-84 and 85-older years of age.**

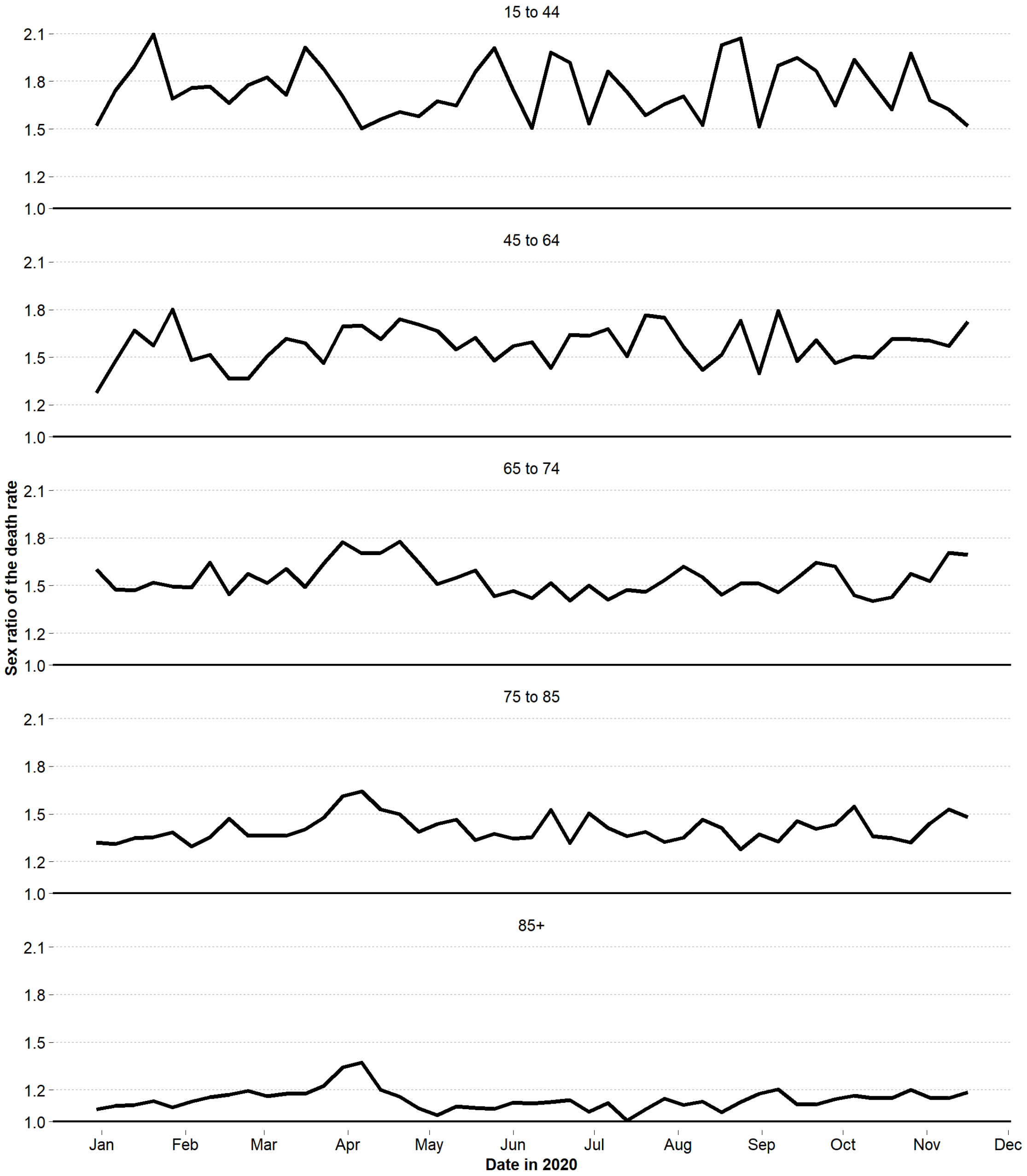
